# Prevalence and Outcomes Associated with Idarucizumab Administration in Trauma Patients on Preinjury Dabigatran Therapy: Analyzing Clinical Utilization in 942 Patients from 74 Hospitals

**DOI:** 10.1101/2024.01.12.24301126

**Authors:** Rebecca L. Moore, Ransom J. Wyse, Justin Jacobs, Samir M. Fakhry, Nina Y. Wilson, Jeneva M. Garland

**Affiliations:** HCA Healthcare/University of Tennessee College of Pharmacy PGY2 Residency Program, Clinical Services Group, HCA Healthcare, Nashville, TN 37203; Center for Trauma and Acute Care Surgery Research, Clinical Services Group, HCA Healthcare, Nashville, TN 37203

**Keywords:** Idarucizumab, Anticoagulant Reversal Agents, Dabigatran, Thromboembolism, Bleeding

## Abstract

**Background:** Increasing numbers of injured patients taking dabigatran are presenting to trauma centers raising an important clinical question: Does reversal with idarucizumab outweigh potential bleeding risks associated with dabigatran? The purpose of this study was to describe the prevalence of idarucizumab administration in trauma patients and compare outcomes for those who received reversal to those who did not.

**Methods:** This retrospective cohort study included trauma inpatients ≥18 years on preinjury dabigatran. Patients were sourced from the registries of Level I–IV trauma centers with an arrival date 1/2017–12/2021. Preinjury dabigatran therapy and idarucizumab administration were confirmed via EMR chart review. Patients on preinjury dabigatran were grouped according to administration status of idarucizumab.

**Results:** 942 trauma patients on preinjury dabigatran (49.7% male; mean GCS:15; ≥70 years: 85.7%) were included, with 10.8% patients reversed with idarucizumab. No statistically significant differences were found for preinjury dabigatran dose (p=0.703), age (p=0.494), blunt injury type (p=0.070), or mechanism of injury (p=0.248). Those reversed with idarucizumab had a greater median head AIS score (3 vs 2; p<0.001), higher proportion full trauma activations (16.7 vs 8.7%; p=0.019), higher median ISS (10 vs 9; p<0.001), were more likely to have a GCS 3–8 (4.9% vs 0.8%; p=0.006), and had increased rates of blood transfusion (4.9% vs 1.3%; p=0.022), ventilator use (10.8% vs 4.7%,p=0.009), and mortality (expired+hospice) (10.8% vs 4.9%; p=0.021). There was no difference between groups for thromboembolic events (1.0% vs 0.7%; p=0.553), hospital LOS (3 vs 4 days; p=0.147), or ICU LOS (3 vs 3 days; p=0.714).

**Conclusions:** In this large, retrospective cohort study of trauma patients, only 10.8% had reversal with idarucizumab. Patients reversed were more severely injured, with increased ICU and ventilator use, more transfusions ≤24 hours, and had increased mortality compared to those not reversed. There was no difference in thromboembolic events between groups. These findings suggest clinicians may be administering idarucizumab based on injury severity *–* especially head trauma *–* regardless of other variables, such as comorbidities. Additional research is needed to determine the optimal role of reversal with idarucizumab vs. other strategies for injured patients on dabigatran.

**Clinical Perspective:** *What is new?:*

- Trauma patients on preinjury dabigatran reversed with idarucizumab were more severely injured and utilized more hospital resources (including increased intensive care unit length of stay and ventilator days) than those not reversed.
- There was no difference in thromboembolic events recorded between trauma patients administered idarucizumab compared to those who were not; however, reversal with idarucizumab was associated with increased rates of blood transfusion and total mortality (expired + hospice).

*What are the clinical implications?:*

- These findings suggest clinicians may be administering idarucizumab to trauma patients based primarily on injury severity, especially head trauma, regardless of other variables, such as comorbidities.
- A large, prospective randomized study of trauma inpatients on preinjury dabigatran that compares dabigatran reversal with idarucizumab to non-specific reversal agents is warranted to establish appropriate criteria for utilization of idarucizumab.

## INTRODUCTION

Warfarin has long been considered the gold standard for anticoagulation for a wide range of conditions. As evidence accumulates on the role of direct oral anticoagulants (DOAC) in the prevention of thrombosis, more patients are transitioning from warfarin to DOAC therapy. From 2011 to 2017, prescriptions for DOACs as a proportion of all oral anti-coagulants increased from 22.1% to 87.3%, whereas warfarin prescriptions decreased from 77.9% to 12.7%.^1^ As expected, with the increase in DOAC prescriptions, there has been an increase in the prevalence of injured patients arriving at hospitals who are on preinjury DOAC therapy, including dabigatran.

Dabigatran is a DOAC approved by the Food and Drug Administration (FDA) to reduce the risk of stroke and systemic embolism in those with atrial fibrillation, to treat deep venous thromboembolism (DVT) and pulmonary embolism (PE) in patients who have been on a 5 – 10 day course of a parenteral anticoagulant, to reduce recurrence of DVT and PE, and for DVT and PE prophylaxis in patients who have recently experienced hip replacement surgery.^2^ Dabigatran is the third most frequently prescribed DOAC– with the majority of patients prescribed the drug being in the geriatric age group.^3^

The proportion of individuals aged ≥65 years in the US is growing, having increased by one-third between 2010 and 2020,^4,5^ and making up 50.7% of all trauma admissions in 2016.^6^ Indications for dabigatran are more prevalent in the geriatric age group (e.g. atrial fibrillation), and as the use of dabigatran and other DOACs has also increased, more trauma patients are presenting on preinjury DOAC therapy. Furthermore, older individuals have an increased tendency to fall, and anticoagulant therapy such as dabigatran, places them at an increased risk of hemorrhage – even life-threatening bleeding – in situations such as traumatic brain injury (TBI).^2,7^ Growth in the geriatric population and increased prescribing of dabigatran places more individuals at risk for bleeding complications from trauma, and underscores the importance of clinical-decision making in the evaluation and management of a trauma patient on preinjury dabigatran, balancing the risks of bleeding against those of thrombo-embolic events.

Idarucizumab is a human monoclonal antibody fragment approved by the FDA for the reversal of dabigatran, specifically binding to the DOAC to neutralize its anticoagulant effect. Importantly, idarucizumab is FDA approved for use in emergency surgeries or urgent procedures and life-threatening or uncontrolled bleeding.^8^ Regardless of these approved indications, the administration of idarucizumab is widely disputed among emergency clinicians due to potential adverse events, cost, and lack of supporting literature concerning its use in trauma patients.^9,10^ Post-marketing and case reports have indicated idarucizumab administration to potentially be associated with an increased thromboembolic risk, including DVT, PE, atrial thrombus, non-ST elevation myocardial infarction (NSTEMI), and ischemic stroke.^11^ Despite reports of these events occurring in less than 1% of patients,^11^ these thromboembolic adverse reactions prompt concern. As trauma patients are inherently in a hypercoagulable state,^12^ clinicians are faced with the decision as to whether reversal with idarucizumab in this patient population outweighs the potential bleeding risks associated with dabigatran.

The purpose of this retrospective cohort study was to describe the prevalence of idarucizumab administration in a large sample of trauma patients from a single, nationwide hospital system and compare the outcomes of trauma inpatients on preinjury dabigatran who received reversal with idarucizumab to those who did not receive idarucizumab.

## METHODS

### Study sample and data sources

In this retrospective cohort study, trauma patients from a large hospital network were included if they were ≥18 years, on pre-injury dabigatran therapy, and presented to the emergency department (ED) between January 1, 2017 and December 31, 2021. Patient demographics including age, sex, race, and injury characteristics (injury type and mechanism) were retrieved from the system-wide trauma registry. Mechanism of injury (MOI) for each patient was identified using International Classification of Diseases, Tenth Revision, Clinical Modification (ICD-10-CM) external cause of injury codes. Preinjury dabigatran use was retrieved from the electronic medical record (EMR), which included patient-reported documentation of home medications in patients’ medical history, while idarucizumab dose and administration were sourced from the ED medication administration record (MAR). Preinjury dabigatran therapy and idarucizumab administration were further confirmed via EMR data from the electronic data warehouse (EDW). Each patient record was assessed for dabigatran therapy and idarucizumab administration, identified by both the generic and trade names (Pradaxa^®^ and Praxbind^®^, respectively) via string search. Only patients taking a preinjury dose of dabigatran 75 mg twice daily or 150 mg twice daily were included; patients who reported preinjury dabigatran 110 mg or dabigatran 220 mg were excluded, as this dose is currently only indicated for prophylaxis of DVT and PE following hip replacement surgery.^2^ Patients with nontraumatic ischemic or hemorrhagic strokes that were present on admission were excluded.

### Variable definitions

The primary goal of the study was to characterize the administration of the reversal agent idarucizumab among trauma patients on preinjury dabigatran therapy. Patients on preinjury dabigatran were grouped according to whether or not they were administered the dabigatran reversal agent, idarucizumab. Administration of idarucizumab – including dosage and time of dose – was sourced from drug administration data in the EDW using string search on the generic name and trade name of Praxbind^®^. Any inconsistencies in the documentation of idarucizumab administration were reviewed by a clinical staff pharmacist on the study team to confirm dosing records for accuracy.

Secondary outcomes included patient-related outcomes for trauma patients on preinjury dabigatran who were administered idarucizumab compared to those who were not administered idarucizumab. These patient outcomes included hemorrhagic events, thromboembolic events, discharge disposition, total mortality, and hospital resource utilization. Hemorrhagic events included hemorrhagic stroke or “bleeding”; since this outcome was not consistently captured in the EDW, “bleeding” was defined as the occurrence of a blood transfusion within the first 0 – 4 hours or 5 – 24 hours of admission as a proxy variable, given this temporal parameter was consistently documented in the trauma registry. Thromboembolic events included thromboembolic stroke, DVT, or PE. Both hemorrhagic stroke and thromboembolic stroke events were sourced from the EDW using ICD-10-CM diagnosis codes (Supplemental Table 1). “Total mortality” was defined as the proportion of patients who expired as an inpatient, or were discharged to hospice. Therefore, “Total mortality” was the sum of inpatient deaths (“Expired”) and hospice discharges (“Hospice”) as documented in the trauma registry.^13^ Hospital resource utilization information was also extracted from the trauma registry, and included length of stay (LOS, reported in days), intensive care unit (ICU) utilization, ICU LOS, ventilator utilization, and days on a ventilator.

The R package ‘comorbidity’ was used to compute a van Walraven weighted Elixhauser score.^14^ Since the specific indication for preinjury dabigatran therapy was not consistently available in the EDW or trauma registry, the EDW was searched for the presence of the comorbidity of interest for which dabigatran therapy is indicated, i.e. atrial fibrillation. History of atrial fibrillation, or the presence of atrial fibrillation on arrival using ICD-10-CM diagnosis codes, was deemed the most likely primary reason for dabigatran therapy (Supplemental Table 1).

### Statistical analyses

Descriptive analyses comparing trauma patients who were administered idarucizumab to those who did not receive idarucizumab were performed for patient demographics, diagnoses, and outcomes using Pearson ^2^ tests of association for categorical variables, and Wilcoxon Rank Sum tests for continuous variables. Fisher’s exact test was used for categorical outcomes with fewer than ten results in each category. *P* values less than .05 were considered statistically significant. R software version 4.2.2 was used for all analyses.^15^ The Institutional Review Board (IRB) of the University of Tennessee Health Science Center waived ethical approval for this work and determined it to be exempt from IRB oversight. The Strengthening the Reporting of Observational Studies in Epidemiology (STROBE) guidelines were utilized in the reporting of this research (Supplemental Table 2).^16^ The authors had full access to all data in the study and take responsibility for the integrity of the data and accuracy of the data analysis.

## RESULTS

### Patient characteristics

There were 500LJ339 patients in the trauma registry that presented to one of 74 facilities, which included 8 Level I, 35 Level II, 20 Level III, and 11 Level IV trauma centers, between January 1, 2017, and December 31, 2021. Only 1177 (0.2%) met the aforementioned study criteria for preinjury dabigatran use. Fifty-two patients admitted to facilities that did not have idarucizumab on formulary during the study period were excluded from the analysis. In addition, admissions that did not meet the inclusion criteria were withdrawn (duplicate trauma identifications = 7; under age 18 = 1; patients on hip replacement 110 mg dabigatran dose = 41; nontraumatic stroke present on arrival = 134). After these exclusions, 942 unique patients remained, with 102 (10.8%) who received idarucizumab and 840 (89.2%) who did not (Figure 1).

**Figure 1.**
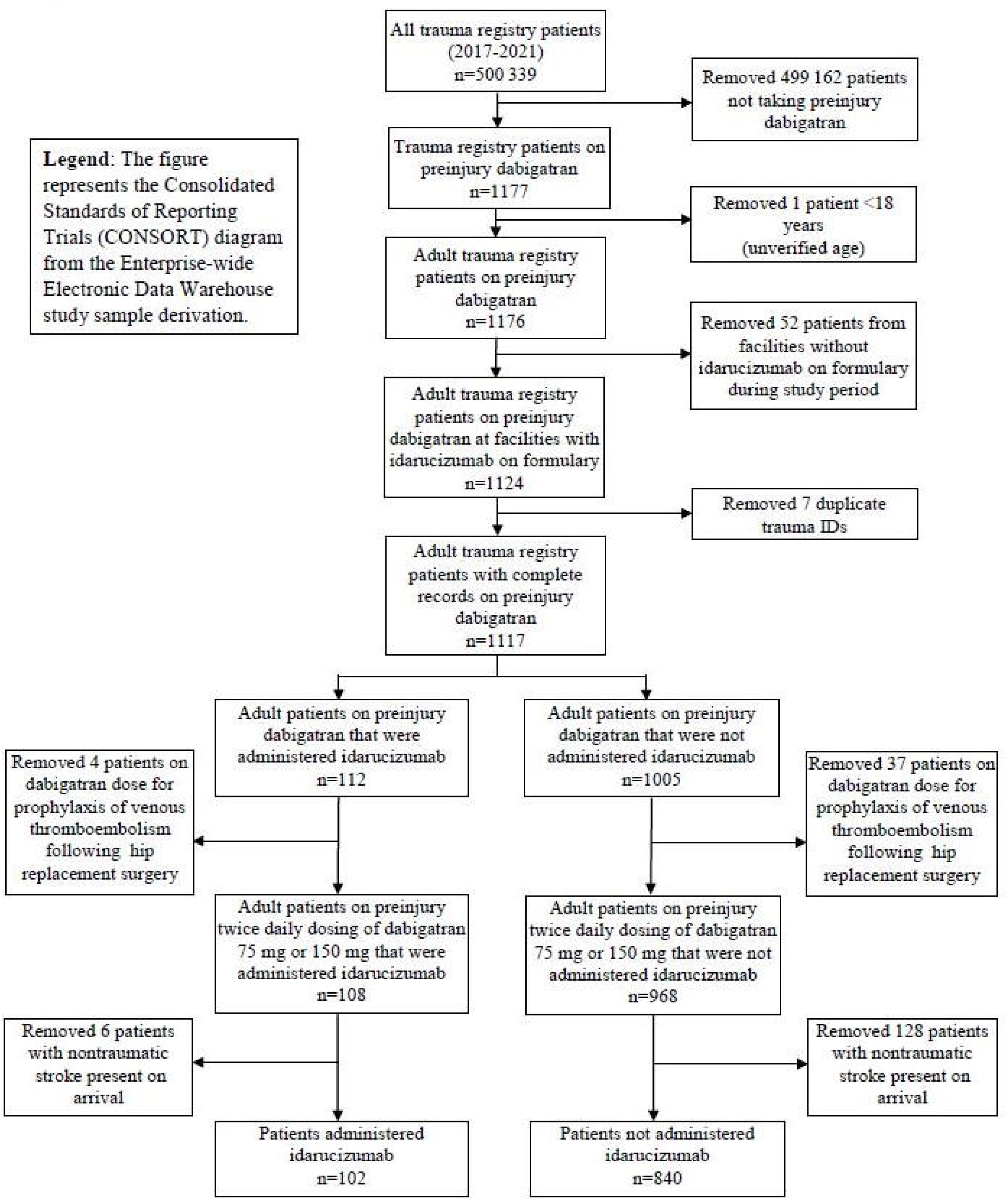
CONSORT diagram for the Comparison of Administered Idarucizumab Study Sample Derivation.

The majority of patients were 70 years of age or older (85.7%), with 484 (51.4%) in the 70 – 84 age range. There were 474 (50.3%) female patients. The study sample was predominantly White (92.9%), with 18 (1.9%) Black patients and 49 (5.2%) Other races. A higher proportion of Black patients were administered idarucizumab compared to those who did not receive idarucizumab (4.9% vs 1.5%, *P*=0.037). Ninety-one patients received the FDA recommended dosage of 5 grams (89.2%), 5 patients received 2.5 grams (4.9%), 2 patients received 10 grams (2.0%), and there was missing dosage administration data for 4 patients (3.9%). There were 795 (84.4%) patients with a documented history of atrial fibrillation, which was identified as a proxy of the indication for their preinjury dabigatran therapy. A statistically significantly larger proportion of patients administered idarucizumab had a full trauma activation on arrival compared to patients who did not receive the reversal drug (16.7% vs 8.7%, *P*=0.019; Table 1). No statistically significant differences were found for sex, age, Elixhauser score, transfers in, indication for dabigatran therapy, or dabigatran dose between patients who were administered idarucizumab and those who were not (Table 1).

**Table 1.** Demographic and Clinical Patient Characteristics.

| Patient Demographics | Overall | Idarucizumab Grouping |  | *P* value * |
| --- | --- | --- | --- | --- |
|  |  | Idarucizumab | No Idarucizumab |  |
|  | N=942 | n=102 (10.8%) | n=840 (89.2%) |  |
| **Female sex, n (%)** | 474 (50.3%) | 47 (46.1%) | 427 (50.8%) | .364 |
| **Race** |  |  |  | .048 |
| White, n (%) | 875 (92.9%) | 90 (88.2%) | 785 (93.5%) | .065 |
| Black, n (%) | 18 (1.9%) | 5 (4.9%) | 13 (1.5%) | .037 |
| Other, n (%) | 49 (5.2%) | 7 (6.9%) | 42 (5.0%) | .475 |
| **Age, y, median [IQR]<sup>†</sup>** | 81 [74, 87] | 80 [74.3, 86.5] | 81 [74, 87] | .650 |
| **Age group** |  |  |  | .494 |
| 18-54 years, n (%) | 19 (2.0%) | 1 (1.0%) | 18 (2.1%) | .711 |
| 55-69 years, n (%) | 116 (12.3%) | 11 (10.8%) | 105 (12.5%) | .750 |
| 70-84 years, n (%) | 484 (51.4%) | 60 (58.8%) | 424 (50.5%) | .117 |
| ≥85 years, n (%) | 323 (34.3%) | 30 (29.4%) | 293 (34.9%) | .320 |
| **Elixhauser score, median [IQR]<sup>†</sup>** | 10 [5, 17] | 9 [5, 16.8] | 11 [5, 17] | .264 |
| **Transfers in, n (%)** | 186 (19.7%) | 20 (19.6%) | 166 (19.6%) | .971 |
| **Activation type** |  |  |  | .010 |
| Full, n (%) | 90 (9.6%) | 17 (16.7%) | 73 (8.7%) | .019 |
| Partial, n (%) | 266 (28.2%) | 31 (30.4%) | 235 (28.0%) | .642 |
| Consult, n (%) | 255 (27.1%) | 30 (29.4%) | 225 (26.8%) | .557 |
| None, n (%) | 266 (28.2%) | 15 (14.7%) | 251 (29.9%) | .001 |
| Unknown/Missing, n (%) | 65 (6.9%) | 9 (8.8%) | 56 (6.7%) | .408 |
| **Preinjury dabigatran indication** |  |  |  | .791 |
| Atrial fibrillation, n (%) | 795 (84.4%) | 87 (85.3%) | 708 (84.3%) | - |
| Other, n (%) | 147 (15.6%) | 15 (14.7%) | 132 (15.7%) | - |
| **Preinjury dabigatran dose** |  |  |  | .703 |
| 75 mg BID<sup>†</sup>, n (%) | 262 (27.8%) | 30 (29.4%) | 232 (27.6%) | - |
| 150 mg BID<sup>†</sup>, n (%) | 680 (72.2%) | 72 (70.6%) | 608 (72.4%) | - |

\*All comparisons between idarucizumab administration and no idarucizumab administration were significant at $P < .05$ .
<sup>†</sup>Abbreviations: IQR=interquartile range; BID=bis in die (twice a day).

### Injury description

Falls were the most common MOI (85.2%), with 639 (67.8%) patients presenting for same-level falls, and 164 (17.4%) categorized in the trauma registry as “other falls” (Table 2). The MOI was not statistically significantly different between those who received idarucizumab and those who did not (*P*=0.248). Overall, median injury severity score (ISS) was 9 [interquartile range (IQR) 4-10], but the group administered idarucizumab had a statistically significantly higher median score than those who did not (10 [IQR 8 – 17] vs 9 [IQR 4 – 9]; *P*<0.001). Similarly, when ISS was categorized, there was a greater proportion of patients in the more severely injured “16 – 24” and “≥25” ISS groupings for those who were administered idarucizumab compared to patients who did not receive the drug (29.4% vs 3.8%, and 10.8% vs 2.1%, respectively; *P*<0.001). Median Glasgow Coma Score (GCS) was 15 [IQR 15 – 15] across the sample, as well as in each group; however, when GCS was examined by category, the idarucizumab group had a statistically significantly higher proportion of patients in the most severe “3 – 8” range, with 5 (4.9%) patients compared to 7 (0.8%) patients who did not receive idarucizumab (*P*<0.001). Furthermore, patients administered idarucizumab were more likely to have a higher head abbreviated injury score (AIS), with the median being 3 [IQR 3 – 4], compared to those who were not administered idarucizumab (2 [IQR 2 – 3]). All injury characteristics are reported in Table 2.

**Table 2.** Injury Patterns of Patients Administered Idarucizumab Compared to Patients Who Did Not Receive Idarucizumab.

|  | Overall | Idarucizumab Grouping |  | *P*<br>value* |
| --- | --- | --- | --- | --- |
|  |  | Idarucizumab<br>n=102 (10.8%) | No Idarucizumab<br>n=840 (89.2%) |  |
|  | N=942 |  |  |  |
| **Injury type** |  |  |  | .070 |
| Blunt, n (%) | 924 (98.1%) | 98 (96.1%) | 826 (98.3%) | .121 |
| Penetrating, n (%) | 8 (0.8%) | 3 (2.9%) | 5 (0.6%) | .046 |
| Other/unspecified, n (%) | 3 (0.3%) | 0 (0.0%) | 3 (0.4%) | 1.000 |
| Missing injury type, n (%) | 7 (0.7%) | 1 (1.0%) | 6 (0.7%) | .553 |
| **MOI** |  |  |  | .248 |
| Same level fall, n (%) | 639 (67.8%) | 63 (61.8%) | 576 (68.6%) | .178 |
| Other fall, n (%) | 164 (17.4%) | 21 (20.6%) | 143 (17.0%) | .406 |
| MVC, n (%) | 59 (6.3%) | 6 (5.9%) | 53 (6.3%) | 1.000 |
| Motorcycle, n (%) | 9 (1%) | 1 (1%) | 8 (1%) | 1.000 |
| Pedestrian, n (%) | 12 (1.3%) | 3 (2.9%) | 9 (1.1%) | .132 |
| Pedal cyclist, n (%) | 10 (1.1%) | 2 (2.0%) | 8 (1.0%) | .296 |
| Assault, n (%) | 7 (0.7%) | 2 (2.0%) | 5 (0.6%) | .170 |
| Other, n (%) | 39 (4.1%) | 4 (3.9%) | 35 (4.2%) | 1.000 |
| Missing MOI, n (%) | 3 (0.3%) | 0 (0.0%) | 3 (0.4%) | 1.000 |
| **ISS, median [IQR]†** | 9 [4, 10] | 10 [8, 17] | 9[4, 9] | <.001 |
| ISS 1 – 8, n (%) | 426 (45.2%) | 27 (26.5%) | 399 (47.5%) | <.001 |
| ISS 9 – 15, n (%) | 422 (44.8%) | 33 (32.4%) | 389 (46.3%) | .008 |
| ISS 16 – 24, n (%) | 62 (6.6%) | 30 (29.4%) | 32 (3.8%) | <.001 |
| ISS ≥25, n (%) | 29 (3.1%) | 11 (10.8%) | 18 (2.1%) | <.001 |
| Missing, n (%) | 3 (0.3%) | 1 (1.0%) | 2 (0.2%) | .291 |
| **AIS** |  |  |  |  |
| Head, median [IQR]† | 3 [2, 3] | 3 [3, 4] | 2 [2, 3] | <.001 |
| Face, median [IQR]† | 3 [2, 3] | 2.5 [2, 3] | 3 [2, 3] | .639 |
| Chest, median [IQR]† | 2 [1, 2] | 2 [1, 2] | 2 [1, 2] | .879 |
| Abdomen, median [IQR]† | 3 [2, 3] | 3 [2, 3] | 3 [2, 3] | .613 |
| Extremities, median [IQR]† | 2 [2, 2] | 2 [2, 2] | 2 [2, 2] | .790 |
| External, median [IQR]† | 1 [1, 1] | 1 [1, 1] | 1 [1, 1] | .268 |
| **GCS, median [IQR]†** | 15 [15, 15] | 15 [15, 15] | 15 [15, 15] | <.001 |
| GCS 3 – 8, n (%) | 12 (1.3%) | 5 (4.9%) | 7 (0.8%) | .006 |
| GCS 9 – 12, n (%) | 16 (1.7%) | 4 (3.9%) | 12 (1.4%) | .085 |
| GCS 13 – 15, n (%) | 859 (91.2%) | 91 (89.2%) | 768 (91.4%) | .459 |
| Missing GCS, n (%) | 55 (5.8%) | 2 (2.0%) | 53 (6.3%) | .113 |

\*All comparisons between idarucizumab administration and no idarucizumab administration were significant at $P < .05$ .
†Abbreviations: MOI=Mechanism of Injury; MVC=motor vehicle collision; ISS=injury severity score; IQR=interquartile range; AIS=Abbreviated injury score; GCS=Glasgow Coma Score.

### Outcomes

In the overall sample, there were 16 (1.7%) patients who received transfusions, indicating significant hemorrhage. Patients administered idarucizumab were more likely to experience bleeding that required transfusion compared to those who did not receive idarucizumab (4.9% vs 1.3%, respectively; *P*=0.022; Table 3).

**Table 3.**
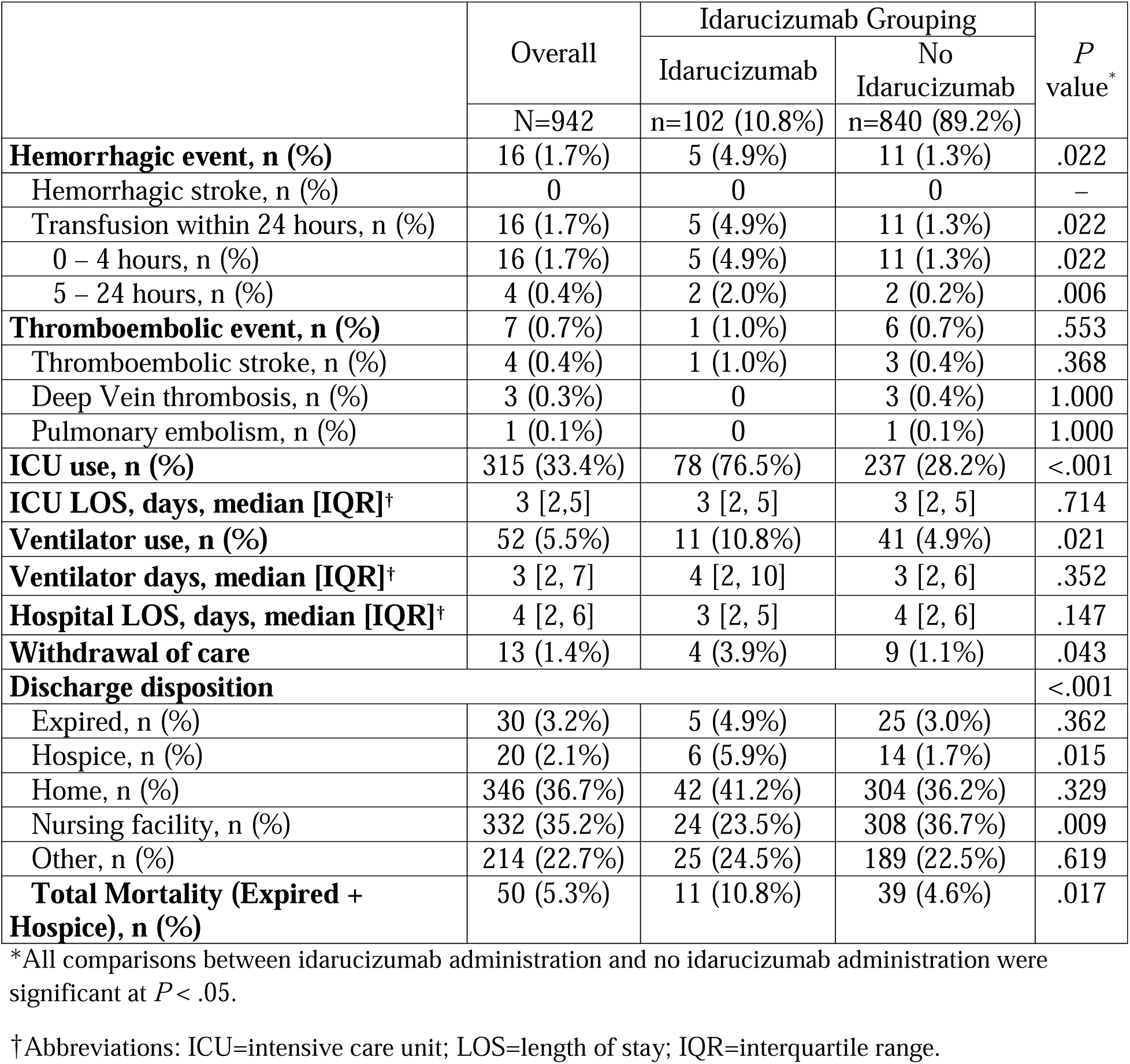
Outcomes of Patients Administered Idarucizumab Compared to Patients Who Did Not Receive Idarucizumab.

A total of 7 (0.7%) patients had a thromboembolic event reported. There were 4 (0.4%) patients who had a thromboembolic stroke, 3 (0.3%) who had a clinically detectable DVT, and 1 (0.1%) who had a PE. The four patients who had either a clinically detectable DVT or PE were in the group that did not receive idarucizumab, but there was no statistically significant difference between the two groups for thromboembolic events of any kind (Table 3).

In terms of hospital resource utilization, 76.5% of those administered idarucizumab had any LOS in the ICU, while 28.2% of those not administered idarucizumab had any LOS in the ICU (*P*<0.001). Despite this difference, patients had a similar median ICU LOS of 3 [IQR 2 – 5] days both across the sample and within groups. Ventilator use was also more prevalent in those administered idarucizumab (11 patients, 10.8%), compared to 41 (4.9%) patients for those who did not receive idarucizumab (*P*<0.021), though the number of days on a ventilator was not statistically significantly different between the two groups, with an overall median of 3 [IQR 2 – 7] days.

The total mortality, which was defined as the sum of patients who expired in the inpatient setting and those who were discharged to hospice, was found to be statistically significantly different between the two groups, with a higher proportion among those administered idarucizumab compared to those who were not given the drug (10.8% vs 4.6%, *P*=0.017). This statistically significant difference in the proportion of total mortality was most affected by those discharged to hospice. In patients who received idarucizumab, 5.9% were discharged to hospice, while 1.7% were discharged to hospice in the patient group who did not receive idarucizumab (*P*=0.015). The number of patients who expired in the inpatient setting was not statistically significantly different between the groups, 5 (4.9%) in the patient group who received idarucizumab compared to 25 (3.0%) in patients who did not receive idarucizumab (*P*=0.362; Table 3).

## DISCUSSION

This retrospective cohort study was conducted to characterize the prevalence of idarucizumab administration in trauma patients on preinjury dabigatran therapy. In the analysis of 942 trauma patients on preinjury dabigatran therapy (0.2% of all trauma patients seen) from 74 level I–IV trauma centers and non-trauma centers, 102 (10.8%) were administered idarucizumab for the reversal of dabigatran. Patients who were administered idarucizumab were found to be more severely injured with a higher ISS and had an increased proportion of total mortality. Compared to those who did not receive the reversal drug, idarucizumab administration was associated with a higher proportion of full trauma activations, increased ICU and ventilator use, and more transfusions within the first 24 hours of admission. There was not a statistically significant difference between the two groups for the number of individuals who expired in the hospital, but patients who received idarucizumab were more likely to be discharged to hospice, which increased total mortality (expired in-hospital + hospice) in the group that received dabigatran reversal with idarucizumab. There were no statistically significant differences in clinically detectable thromboembolic events – including thromboembolic stroke, DVT, and PE – between those who were administered idarucizumab and those who were not.

As the numbers of elderly patients continues to increase, comorbidities associated with aging – such as atrial fibrillation^17^ – are also on the rise and frequently necessitate the use of anticoagulant medications, including dabigatran.^17-19^ Despite the protective anticoagulative properties dabigatran has to offer patients with atrial fibrillation, its use may pose increased bleeding risks following injury. When emergency surgery or urgent procedures are needed, or a patient presents with life-threatening or uncontrolled bleeding, the reversal of dabigatran may be required. Idarucizumab binds to dabigatran with 350 times greater affinity than does thrombin, thereby negating its anticoagulant effect, which is concerning, as studies have shown increased adverse events in elderly patients when dabigatran is reversed with idarucizumab.^20,21^ Similar to other studies regarding the use of idarucizumab in the reversal of dabigatran, the majority of patients who met eligibility criteria in this study were elderly.^22-24^ Despite the majority of dabigatran reversal with idarucizumab tending to occur in the elderly, this study found there was no association between increasing age and idarucizumab administration among trauma patients.

Several studies have reported the safety of idarucizumab administration. A retrospective case-control study that included hospitalized patients for dabigatran-associated nontraumatic gastrointestinal bleeding or intracranial bleeding by Singh et al noted there were no significant differences in mortality (odds ratio, 1.39 [95% CI, 0.51–3.45]) or venous thromboembolism (odds ratio, 0.35 [95% CI, 0.08–1.58]) for those who received idarucizumab compared to those who did not receive the reversal drug.^25^ Similarly, a meta-analysis by Chaudhary et al stated idarucizumab had a “promising safety profile” with 82% anticoagulation reversal and low mortality (11%) and thromboembolic events (5%).^24^ Chaudhary et al also instill a hint of doubt by stating, “*While the safety data from these landmark trials were promising, head-to-head trials have not compared the safety and outcomes of idarucizumab or andexanet alfa (AA) with traditional nonspecific reversal agents (FFP, 4F-PCC, or A-PCC).”*^24^ Furthermore, the American College of Gastroenterology-Canadian Association of Gastroenterology Clinical Guideline recommends against the use of idarucizumab for those with gastrointestinal bleeding due to limited evidence of benefit and the high cost of the drug.^26^

In this study, a statistically and clinically significant proportion of patients who received idarucizumab were more likely to be severely injured and have a full trauma activation, compared to those who did not. Patients administered idarucizumab were more likely to have a higher ISS, a higher rate of transfusion, a higher AIS of the head, a lower GCS, and increased ICU and ventilator use. Consistent with these markers of severity and increased risk, they were also more likely to have higher total mortality (expired in hospital + hospice). This statistically significant increased mortality for those who received idarucizumab is more likely related to the patient’s injury severity than to the administration of idarucizumab. Interestingly, compared to those who did not receive idarucizumab, patients who were administered the drug were less likely to expire in the hospital, yet the total mortality calculation (expired in hospital+ hospice) indicated a statistically significant increase in total mortality for those administered idarucizumab, suggesting discharge to hospice – and not age itself as reported above – may play a role in this outcome as reported in previous studies of trauma patients.^27,13^

Regardless of any hesitancy to utilize the drug, adherence to the idarucizumab FDA recommended dosing appears to be relatively high. There are few large studies concerning idarucizumab dosage compliance. Our study – with 89.2% compliance – was similar to a small study by Mitrovic, et al. that included 68 patients on pre-admission dabigatran.^22^ They reported 94% of patients were administered idarucizumab with the appropriate FDA dosing guidance.

Idarucizumab administration may place trauma patients at an increased risk of thromboembolic events, not only due to the coagulative drug properties of idarucizumab binding to dabigatran and negating its anticoagulative effects, but also due to the fact that trauma patients present in a hypercoagulable state.^28^ This study found no statistically significant differences in clinically detectable thromboembolic events between groups. All patients were not actively surveilled for this outcome or evaluated for these potential adverse events following discharge, limiting the conclusion that may be drawn from this finding.

The clinically important findings reported in this study may be indicative of specific effects of idarucizumab in the trauma patient, as few studies have been conducted strictly in this high-risk population. A small study of 15 trauma patients by Oberladstätter et al found similar results to this study, in which no patients who received idarucizumab experienced clinically detectable thromboembolic events. Conversely, previous studies that included non-trauma patients found idarucizumab administration to be associated with increased clinically detectable thromboembolic events.^20,29^

### Limitations

The usual cautions regarding retrospective analysis of administrative datasets are warranted, including the relatively small sample size. Due to the nature of patient-reported preinjury dabigatran use, there is the potential for cognitive bias. Therefore, we could not verify the time of the patients’ last dose of dabigatran with certainty. Our dataset was also lacking detailed information concerning the indication for idarucizumab administration, as well as outcomes associated with each idarucizumab dose (eg, the cause of death or whether there was any expansion of intracranial hemorrhages present on admission). Furthermore, initial dabigatran serum concentration values and dabigatran serum concentration values following administration of idarucizumab were not available.

### Conclusions

This retrospective cohort study in trauma patients found 983 patients (0.2%) from 87 level I–IV trauma centers were on preinjury dabigatran therapy and 102 (10.4%) were reversed with idarucizumab. Patients administered idarucizumab were found to be more severely injured, and had increased ICU and ventilator use, more transfusions within the first 24 hours of admission, and an increased total mortality compared to those who did not receive idarucizumab. There was no statistically significant difference in clinically detectable thromboembolic events between the two groups. These findings suggest clinicians may be administering idarucizumab based primarily on injury severity, especially head trauma, regardless of other variables such as comorbidities. There may be other more clinically appropriate dabigatran reversal agents in trauma, which warrants further investigation. Additional research is needed to determine the optimal role of reversal with idarucizumab vs. other strategies for injured patients on dabigatran.

## Data Availability

The authors had full access to all data in the study and take responsibility for the integrity of the data and accuracy of the data analysis.

## NON-STANDARD ABBREVIATIONS AND ACRONYMS

DOAC: direct oral anticoagulants
FDA: Food and Drug Administration
DVT: deep vein thrombosis
PE: pulmonary embolism
TBI: traumatic brain injury
NSTEMI: non-ST elevation myocardial infarction
ED: emergency department
MOI: mechanism of injury
ICD-10-CM: International Classification of Diseases, Tenth Revision, Clinical Modification
EMR: electronic medical record
MAR: medication administration record
EDW: electronic data warehouse
ICU: intensive care unit
LOS: length of stay, reported in days
IRB: institutional review board
STROBE: Strengthening the Reporting of Observational Studies in Epidemiology
ISS: injury severity score
IQR: interquartile range
GCS: Glasgow Coma Score
AIS: abbreviated injury score

## ACKNOWLEDGMENTS, SOURCES OF FUNDING, & DISCLOSURES

### Acknowledgments

None

## SUPPLEMENTAL MATERIAL

Tables S1-S2

## SUPPLEMENTAL FILES

Supplemental files are intended for publication as an online data supplement.

